# The impact of systematized generation, evaluation, and incorporation of machine learning algorithms for clinical variant classification

**DOI:** 10.1101/2025.02.03.25321356

**Authors:** Laure Fresard, Flavia M Facio, Elaine Chen, Alexandre Colavin, Britt Johnson, Carlos Araya, Toby Manders, Alexander Wahl, Hillery Metz, Jack Nicoludis, Karen Ouyang, Samskruthi Padigepati, Yuya Kobayashi, Jason Reuter, Keith Nykamp

## Abstract

Variants of uncertain significance (VUS) pose a significant challenge for those undergoing genetic testing, leading to prolonged uncertainty and inappropriate medical care. VUS rate reduction is critical to fully realize the utility of genetic testing for all populations. With the growth of large-scale biological data sources and modern Machine Learning (ML) techniques, predictive modeling has enormous potential for VUS reduction.

For this purpose, we developed the Invitae Evidence Modeling™ Platform (EMP), with key features designed to maximize the utility and confidence of predictive algorithms for variant classification. First, input data for a new model is curated to correspond to a single major evidence category within a variant classification framework. Second, gene-specific training and/or validation is performed for each model type. Third, accuracy thresholds are set to filter out gene-specific models that do not meet stringent accuracy metrics. Finally, prediction scores for variant pathogenicity are calibrated to ensure internally consistent evidence weighting within the classification framework.

The EMP has accelerated the development of ML algorithms and greatly expanded the amount of evidence available for variant classification. EMP evidence has been applied to more than 800,000 variants across 1 million individuals, 42% of which would have been VUS without this evidence. Importantly, definitive classifications (P, LP, LB, B) made with EMP evidence have high prospective concordance (>99%) with ClinVar submissions. Finally, we demonstrate that further use and development of EMP evidence for variant classification has the potential to reduce the VUS disparity across race/ethnicity/ancestry (REA) groups.

## Introduction

The promise of the Human Genome Project ^1,2^ and clinical genomics ^3,4^ is to provide personalized and actionable insights for healthcare, including screening recommendations, reproductive guidance, and treatment decisions. While tremendous progress towards this promise has been made for many disease areas ^5^, one of the biggest remaining challenges is our limited understanding of genetic variation throughout the genome. Clinical variant classification is the process of determining whether a DNA sequence variant is likely to increase the risk for a given disease. To establish risk, it is critical to demonstrate a strong correlation between one or more genetic variants and a clinical phenotype. However, the majority of variants detected in the population and observed in individuals undergoing genetic analysis are exceedingly rare (observed less than 1 in 10,000 people sequenced).^6–9^ Compounding this challenge, potential disease-causing variants must be distinguished from millions of rare neutral or benign variants found in an individual. Given these challenges, classical methods employing odds ratios and case-control studies have limited utility for high-throughput clinical variant classification. Instead, robust methods for this problem make use of multiple lines of orthogonal evidence, which individually may be limited or weak but when combined can provide sufficient confidence that a variant is likely to be associated with disease.

The American College of Medical Genetics (ACMG), together with the Association for Molecular Pathology (AMP), have provided standard terminology (pathogenic, likely pathogenic, uncertain significance, likely benign, benign) for describing an association of a DNA sequence variant with a Mendelian disease, and guidance for the types of data (e.g., population, computational and predictive, functional, segregation, de novo, allelic phase) that are useful for clinical variant classification.^10^ In addition, ClinVar was established through the National Institutes of Health (NIH) to provide a centralized public database for sharing variant classifications and supporting evidence from the clinical genetic and research communities.^11,12^ To date, variant classification submissions have been posted for > 3 million unique germline variants from more than 2500 organizations, including most major clinical testing labs in the US (https://www.ncbi.nlm.nih.gov/clinvar/, accessed October 2024). ClinVar provides a rich source of information for comparing variant classifications across multiple genetic testing laboratories, identifying trends in classification rates, quality, and consistency, and following the growing knowledge about variants and their classifications over time.

A key takeaway from ClinVar is that variants of uncertain significance (VUS) are a significant and growing challenge for testing laboratories and those undergoing genetic testing.^13^ Just over 52% of all unique variants in ClinVar have either a VUS classification or conflicting submissions (https://www.ncbi.nlm.nih.gov/clinvar/, accessed October 2024), which translates into at least one VUS finding for ∼30-40% of all clinical reports delivered from commercial laboratories in recent years.^14,15^ Importantly, a VUS report can lead to feelings of frustration with genetic testing, worry about future changes to management recommendations due to reclassifications, prolonged uncertainty, and lack of timely and appropriate medical care.^16–18^ As a result, strategies to reduce VUS in a timely manner are of utmost importance in clinical genetic testing.

With this in mind, we developed a framework for developing, validating and integrating machine learning (ML) evidence, described as the Evidence Modeling Platform (EMP), designed to aggregate and generate new types of evidence specifically for clinical variant classification across all major ACMG evidence categories. ML techniques have a significant advantage in their ability to detect complex patterns in large genomic datasets, which can be used to predict the pathogenicity of millions of genomic variants. Since these patterns are often beyond what a human can observe, ML predictions have the power to dramatically reduce VUS rates by unlocking additional evidence from data that is inefficiently utilized by traditional variant classification frameworks and dramatically reduce VUS rates.

Due to significant advances in ML methods and the proliferation of high quality classification data, we’ve seen a proliferation of highly sophisticated prediction models for variant classification in recent years.^19–26^ While this is a sign of great progress in the field of computational biology and genomics, it creates several challenges for large-scale testing laboratories and individual practitioners of variant classification alike. First, with so many high-performing models to choose from, it can be difficult to decide which predictions to use for each variant. Second, once a model has been chosen, the prediction scoring metrics need to be calibrated to evidence strength within the variant classification framework of choice. Third, it can be difficult to know which evidence criteria can be applied independently of a prediction model because the underlying data used to train the model may encompass multiple evidence categories, including population data, clinical data, experimental and/or *in silico* data. Finally, addressing these challenges is time consuming and difficult to do across a large set of genes.

In this report, we present the components of the EMP (Figure 1) designed to address these challenges and maximize the utility of ML approaches for clinical variant classification. We demonstrate that systematizing the development, validation and calibration of pathogenicity prediction tools greatly increases the amount of evidence that is useful for variant classification in more than 1 million individuals tested for genetic diseases. Finally, we discuss the impact of implementing a robust and scalable evidence modeling program on VUS rates across a wide range of clinical areas and VUS disparity across race/ethnicity/ancestry (REA) groups.

**Figure 1:**
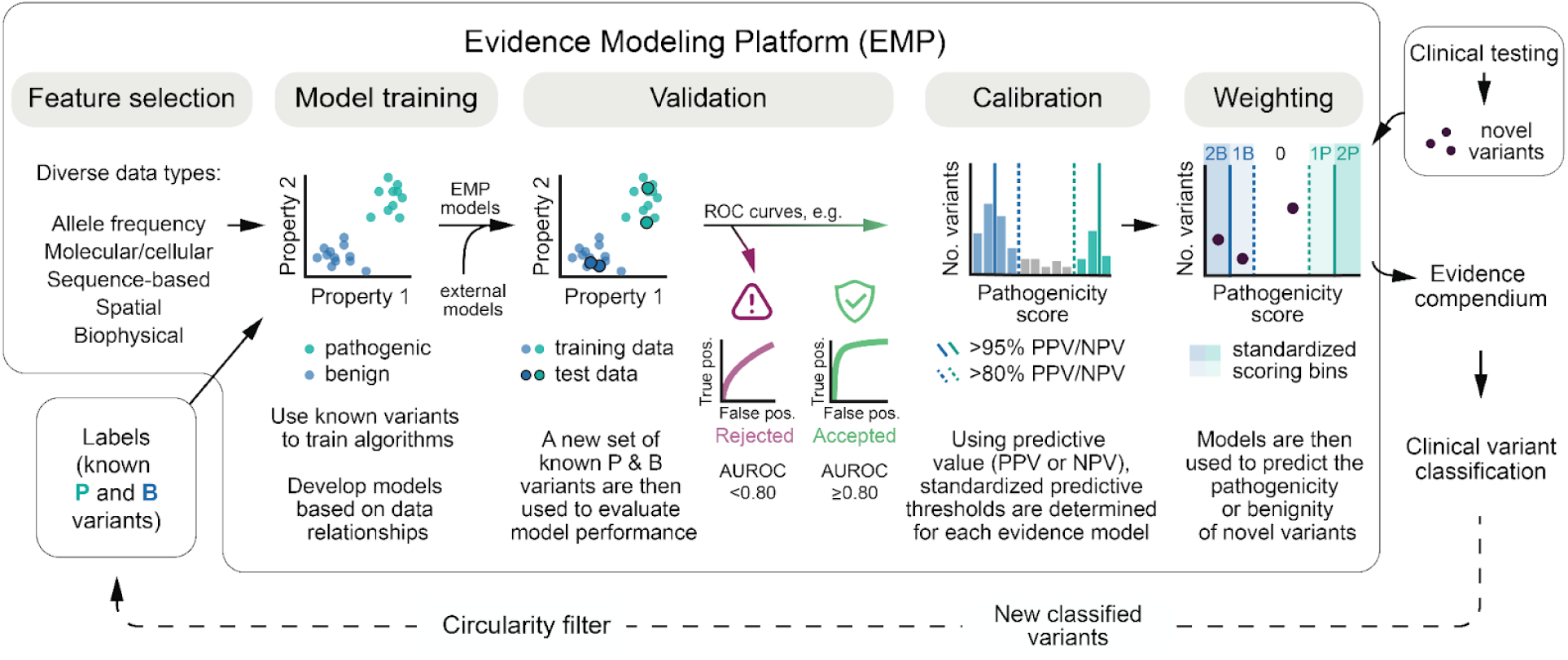
Evidence Modeling Platform pipeline. Schematic illustrating the general pipeline used to generate, evaluate, and integrate modeling evidence into clinical variant classification. Note that while only two properties (Property 1 and Property 2) are depicted (for simplicity), evidence models typically have high dimensionality with many properties of a variant being used for training. A wide range of discrete machine learning (ML) models/algorithms (see methods) are used to examine the many attributes of a gene variant that may or may not contribute evidence of its pathogenicity, such as allele frequency, sequence evolution, protein stability, structural changes and functional data. EMP outputs pathogenicity scores that are calibrated, weighted and included in an evidence compendium. Clinical experts then use this compendium to classify genetic variants following ACMG guidelines. Variants classified using specific types of EMP evidence are excluded from subsequent rounds of model training to avoid circularity bias and inflated performance estimates for newly trained models.

## Subjects and methods

### Clinical cohort and genetic testing

DNA sequencing results from unrelated individuals (N=3,445,847) referred for genetic testing at Invitae^®^ (now Labcorp Genetics) between September 1, 2013 and October 1, 2023 were included in this study. Individuals who requested data deletion during this time period were not included. Next-generation sequencing was performed as previously described.^27–29^ The use of de-identified patient data for this study was approved by an independent Institutional Review Board (approval number 20161796; WCG).

Information on patient age, gender, and race/ethnicity/ancestry (REA) was provided by ordering clinicians. Clinicians selected from available choices for race and ethnicity on test requisition forms: Ashkenazi Jewish, Asian, Black, French Canadian, Hispanic, Native American, Pacific Islander, Sephardic Jewish, White (non-French Canadian, non-Hispanic, and non-Jewish for the purposes of this study), or Other. When Other was selected, a free-text response could be added. If free-text responses matched a preset group, individuals were included in that group.

Individuals with more than one reported REA group were grouped as multiple.

### Variant classification

Variants were classified as benign (B), likely benign (LB), variants of uncertain significance (VUS), likely pathogenic (LP), or pathogenic (P) using Sherloc,^30^ a validated variant classification framework based on guidelines from the American College of Medical Genetics and Genomics and the Association for Molecular Pathology.^10^ Sherloc is a framework for matching variant data (i.e., allele frequency, case reports, family segregation, assay data, computational prediction scores) to predefined evidence criteria. Each predefined evidence criterion is assigned a score, in half-point increments, that range from 0.5 - 5.0 pathogenic points (PP) or from 0.5 - 5.0 benign points (BP). A preliminary classification is determined by summing PP and BP for each variant independently. The threshold for P is ≥ 5PP, LP is ≥4PP *and* <5PP, LB is ≥3BP *and* <5BP, B is ≥5BP. Variants with <4PP *and* <3BP are generally classified as VUS. Trained variant scientists and/or board-certified lab directors review all available evidence and confirm or modify final classifications as appropriate. Variants with both ≥4PP *and* ≥3BP may be classified as VUS, LP/P or LB/B depending on the strength of the conflicting evidence and other data not initially considered during the application of evidence criteria.

### Evidence Modeling Platform (EMP) framework

The steps involved in developing a new model within the EMP framework are illustrated in Figure 1 and described in more detail below.

#### Feature selection

First, data sources, or features, must be selected for model training. To assist with validation and downstream integration of model predictions for clinical variant classification, we selected features for each new model that correspond to a single evidence category as defined by Sherloc.^30^ In this study, features were used that mapped to the following four major evidence categories: (1) Population Data, (2) Computational: Splicing Effect, (3) Computational: Protein Effect or (4) Experimental: Protein Effect (see Table 1 for a set of models presented to illustrate the platform).

**Table 1.** Compendium of evidence models available for clinical variant classification. Each row describes a single evidence model developed with the EMP framework. Columns provide the model name and abbreviation used throughout the manuscript, the corresponding evidence category for which the model maps for variant classification, the types of variants for which the model makes predictions, features used to train the model, data sources for the features, the total number of genes included in the training dataset and the number of genes with at least one variant prediction that passed validation and calibration steps.

| Model Name (Abbreviation) | Evidence Category | Variant type | Features | Data Sources | # Genes trained | # Genes Passing Calibration |
| --- | --- | --- | --- | --- | --- | --- |
| Population Frequency Model (PFM) | Population Data | Nonsense, Missense, Synonymous | Allele Frequency, Gene constraint, Local constraint | gnomAD v2.1.1<sup>31</sup> | 823 | 591 |
| SpliceAI | Computational: Splicing Effect | Splicing | Genomic sequence of pre-mRNA transcripts | GENCODE Annotations<sup>32</sup><br>https://github.com/invitae/SpliceAI | N/A | N/A |
| Gene-Specific Engine (GSE) | Computational: Protein Effect | Missense | Sequence coevolution, physico-chemical properties, protein conservation metrics | Invitae derived (Supplemental Methods) | 957 | 664 |
| Gene-Specific Evolutionary Variant Model (gsEVE) | Computational: Protein Effect | Missense | Evolutionary variant effect prediction scores | Multiple sequence alignments from UniRef100<sup>33</sup> | 396 | 200 |
| Multi-Gene Evolutionary Variant model (mgEVE) | Computational: Protein Effect | Missense | Evolutionary variant effect prediction scores | Multiple sequence alignments from UniRef100<sup>33</sup> | 3333 | 3039 |
| Molecular Stability Engine (MSE) | Computational: Protein Effect | Missense | Predicted change in stability from FoldX calculations ($\Delta\Delta G$) | Invitae derived (Supplemental Methods) | 583 | 234 |
| Deep Mutational Assays (DMA) | Experimental: Protein Effect | Missense | Multiplexed Assays of variant effects (MAVEs) scores | BRCA1<sup>34</sup>, BRCA2<sup>35</sup>, MSH2<sup>36</sup>, SCN5A<sup>37</sup> | 11 | 4 |
| Combined Mutational Assays (DMX) | Experimental: Protein Effect | Missense | Functional Assay results | TP53<sup>38-40</sup> | 3 | 1 |
| Deep Mutational Learning (DML) | Experimental: Protein Effect | Missense | Single-cell mRNA expression | Invitae derived<sup>41</sup> | 40 | 19 |

#### Model Training

In the second step, standard machine learning approaches are used to develop predictive models with the features as input. When using a supervised learning approach, variants of known significance (i.e., pathogenic or benign) are used as labels. For new models described in this study (Table 1), known variants were curated from the Invitae database of classified variants and from ClinVar submissions from laboratories with vetted assertion criteria for variant classification; labs were excluded if no assertion criteria were provided or if lab-specific criteria were inconsistent with ACMG/AMP guidelines. Pathogenic (P) and likely pathogenic (LP) variants were included as pathogenic labels; benign (B) and likely benign (LB) variants were included as benign labels. Variants with high population allele frequencies (> 0.1% in gnomAD) were also included as benign labels. For each model, labels were computed at either the DNA (HGVS.g) or protein (HGVS.p) level to match the type of features used for training.

With the EMP, both gene-specific (gs) and multi-gene (mg) models can be trained. In general, gene-specific training is preferred. When training Population Frequency Models (PFM), gene-specific engines (GSE), gene-specific EVEs (gsEVE), Deep mutational assays (DMA), combined mutational assays (DMX), and deep mutational learning (DML) models, we included all genes for which there were ≥8 pathogenic and ≥8 benign labels, while training of molecular stability engine (MSE) models was limited to genes with ≥5 pathogenic and ≥5 benign labels. For all gene-specific models training was performed for all annotated transcripts (Ensembl, GENCODE v91). The multi-gene EVE (mgEVE) was trained as a single model with 64,304 (pathogenic = 25,855; benign = 38,449) labels collated from 3333 genes. For that later model, labels were stratified by gene such that labels from the same gene were not in different validation splits or in both the training and test splits.

To avoid performance inflation due to circularity and overfitting the data, we filtered labels for training when the definitive classification (P, LP, LB, B) depended on evidence within the same category. For example, if a model was trained with mutational assay data (i.e. MAVEs), we excluded all labels in which the definitive classification depended on data from a MAVE experiment.

The platform employs a systematic approach to identify the optimal ML algorithm and configuration for each trained model. For the models described here, we considered a range of algorithms, including logistic regression, support vector machines (SVM), random forests, and gradient boosting machines (GBM). For hyperparameter optimization, we employed an iterative grid search approach, starting with a wide range of hyperparameter combinations. For both model selection and hyperparameter optimization, we utilized k-fold cross-validation with k set to 5 and 3 repeats. We selected the most regularized and simplest model for each task that achieved performance within measurement error of the best models tried. The selection of the final model and configuration was based on a comprehensive evaluation of cross-validated performance metrics, including accuracy, precision, recall, and F1 score. More details for each of the models described in Table 1 are provided in Supplementary Methods.

#### Model validation

The third step is to determine the accuracy of each independently trained model and filter out those with low performance. Specifically, for the models reported here, the hold-out set of variants from the k-fold validation was used to calculate the area under the receiver operating characteristic (AUROC) for each model, whether gene-specific or multigenic. All models with an AUROC < 0.8 were excluded from subsequent steps. For genes with multiple transcript models with AUROC ≥ 0.8, the transcript model with the highest AUROC was selected for the next step, unless a more clinically relevant transcript was used for clinical reporting purposes.

#### Calibration of prediction scores

Considering the discriminatory power (AUROC) is not consistent across all independently trained models (see Figure S1), the fourth step involves calibration of model prediction scores for use as evidence in variant classification. For this, we developed an approach to calculate the approximate predictive value for each variant score produced by any independently trained model. The calibration step was performed for each gene-specific and multi-genic model with ≥ 5 pathogenic and ≥ 5 benign labels.

Calibration was performed by sorting variant prediction scores for each model from 1 (pathogenic) to 0 (benign). Using all available labels, a positive predictive value (PPV) was calculated for the entire set of variants. Beginning with the lowest (i.e., most benign) prediction score, variants were excluded one at a time until PPV ≥ 95% was obtained. The bin of variants with PPV ≥ 95% was then removed and the PPV was calculated for the remaining variants. If the PPV of this set was < 80%, variants with the lowest scores were again removed until the PPV achieved ≥ 80%. With this analysis, bins of variants corresponding to high (≥ 95%), moderate (≥ 80% and < 95%) and low (< 80%) PPV were created for each model. An example is provided for three genes from the MSE model in Figure S2. This same analysis was repeated for benign prediction scores to establish bins with very high ( ≥ 99%), high (≥ 95% and < 99%), moderate (≥ 80% and < 95%) and low (< 80%) negative predictive values (NPV) for pathogenicity. Note, very high (≥ 99%) NPV bins were only calculated for the population frequency model (PFM).

#### Establishing evidence weight for calibrated predictive value bins

The final step in the EMP framework is to assign calibrated PPV and NPV bins to weighted evidence criteria that will be used for variant classification. Experiments with pre-existing Sherloc evidence criteria demonstrated that 1 point corresponds to ∼80-95% predictive value and 2 points corresponds to ∼95% predictive value (Figure S3). Based on this evidence evaluation, each NPV and PPV bin from the calibration step above is assigned a Sherloc point score as shown in (Figure S4). New Sherloc evidence criteria were created for each model category and predictive value (PV) bin (Table 2).

**Table 2:** Evidence criteria for incorporation of EMP prediction scores within the Sherloc classification framework. Each row represents a single evidence criterion that may be applied within the Sherloc variant classification framework. Columns provide the title and description of the evidence criterion, the major category of evidence for which the criterion maps to within the Sherloc framework, the evidence models that map to the evidence criterion, the benign or point score for each criterion, the number of variants observed in Invitae’s database for which the evidence criterion has been applied, and concordance between the point score direction (pathogenic or benign) and the observed classification for each variant from the previous column.

| Sherlock evidence criterion description [code] | Evidence Category | Model(s) Included | Predictive Value Bin | Benign score | Pathogenic score | Total Variant Count | Invitae Classification Concordance |
| --- | --- | --- | --- | --- | --- | --- | --- |
| Modeling: Population: Very highly predictive benign score [EV0353] | Population Data | PFM | $\geq 99\%$ NPV | 5 | | 2,010 | 98.7% |
| Modeling: Population: Highly predictive benign score [EV0354] | Population Data | PFM | $\geq 95\text{--}99\%$ NPV | 3 | | 15,278 | 99.1% |

| **Sherloc evidence criterion description [code]** | **Evidence Category** | **Model(s) Included** | **Predictive Value Bin** | **Benign score** | **Pathogenic score** | **Total Variant Count** | **Invitae Classification Concordance** |
| --- | --- | --- | --- | --- | --- | --- | --- |
| Modeling: Population: Moderately predictive benign score [EV0355] | Population Data | PFM | ≥80-95% NPV | 1 |  | 17,590 | 96.3% |
| Modeling: Population: Moderately predictive pathogenic score [EV0356] | Population Data | PFM | ≥80% PPV |  | 1 | 4,655 | 97.4% |
| Modeling: SpliceAI: Predicted no impact [EV0361] | Computational: Splice Effect | SpliceAI | N/A | 1 |  | 305,912 | 99.9% |
| Modeling: SpliceAI: Predicted splice site loss [EV0359] | Computational: Splice Effect | SpliceAI | N/A |  | 1 | 23,356 | 95.6% |
| Modeling: SpliceAI: Predicted gain of a de novo splice site [EV0360] | Computational: Splice Effect | SpliceAI | N/A |  | 1 | 14,437 | 90.8% |
| Modeling: Multi-Assay: Very highly predictive benign score [EV0358] | Experimental: Protein Effect | DMX | ≥97.5% NPV | 2.5 |  | 2 | 50.0% |
| Modeling: Multi-Assay: Very highly predictive pathogenic score [EV0352] | Experimental: Protein Effect | DMX | ≥97.5% PPV |  | 2.5 | 152 | 100% |
| Modeling: Assay: Highly predictive benign score [EV0314] | Experimental: Protein Effect | DMA<br>DML | ≥95% NPV | 2 |  | 95 | 90.5% |
| Modeling: Assay: Moderately predictive benign score [EV0313] | Experimental: Protein Effect | DMX<br>DMA<br>DML | ≥80-95% NPV | 1 |  | 343 | 96.5% |
| Modeling: Assay: Highly predictive pathogenic score [EV0310] | Experimental: Protein Effect | DMA<br>DML | ≥95% PPV |  | 2 | 341 | 98.2% |
| Modeling: Assay: Moderately predictive pathogenic score [EV0311] | Experimental: Protein Effect | DMX<br>DMA<br>DML | ≥80-95% PPV |  | 1 | 18 | 88.9% |
| Modeling: InSilico: Highly predictive benign score [EV0319] | Computational: Protein Effect | GSE<br>gsEVE<br>mgEVE<br>MSE | ≥95% NPV | 2 |  | 27,002 | 98.1% |
| Modeling: InSilico: Moderately predictive benign score [EV0318] | Computational: Protein Effect | GSE<br>gsEVE<br>mgEVE<br>MSE | ≥80-95% NPV | 1 |  | 4131 | 71.0% |
| Modeling: InSilico: Highly predictive pathogenic score [EV0315] | Computational: Protein Effect | GSE<br>gsEVE<br>mgEVE<br>MSE | ≥95% PPV |  | 2 | 17,549 | 95.9% |
| Modeling: InSilico: | Computational: | GSE | ≥80-95% |  | 1 | 4925 | 89.9% |

| Sherloc evidence criterion description [code] | Evidence Category | Model(s) Included | Predictive Value Bin | Benign score | Pathogenic score | Total Variant Count | Invitae Classification Concordance |
| --- | --- | --- | --- | --- | --- | --- | --- |
| Moderately predictive pathogenic score [EV0316] | Protein Effect | gsEVE<br>mgEVE<br>MSE | PPV |  |  |  |  |

### Validation and calibration of SpliceAI for variant classification

The SpliceAI model uses transcript sequences to predict the probability of a splicing change given a DNA variant.^32^ The sensitivity of SpliceAI was assessed using a set of 1,236 true positive variants with observed splicing shown through *in vitro* or *in vivo* RNA analysis or *in vitro* splicing assays. SpliceAI Δ scores ≥ 0.2 were used as positive predictions for a splicing change and SpliceAI Δ scores < 0.2 were used as negative predictions for a splicing change by SpliceAI. With this threshold, we observed 91.3% sensitivity (1129/1236 true positives) and 79% specificity (64/81 true negatives) for known DNA variants identified with Invitae’s hereditary cancer splicing assay.^29^ DNA variants predicted to have a splicing impact (Δ scores ≥ 0.2) were given 1 pathogenic point while DNA variants predicted to have no impact on splicing (Δ scores < 0.2) were given 1 benign point (see Table 2). We used a modified version of SpliceAI allowing batch processing of variants in order to generate predictions (https://github.com/invitae/SpliceAI).

### Rules for combining modeling evidence for variant classification

To prevent over-counting evidence from two or more models that share underlying data and features, we established two rules for deciding (1) which models are used for each evidence criteria and (2) which evidence criteria should be counted for variant classification. First, for evidence criteria that may be informed by >1 model (see Table 2, column 3), we selected the gene-specific model (e.g., GSE, gsEVE or MSE) with the highest AUROC. For example, if the gsEVE model was found to have a higher AUROC than GSE or MSE for a given gene, the PPV and NPV bins from the calibration step for gsEVE prediction were used to inform which evidence criteria are assigned to each variant in this gene. If a gene-specific model and multi-gene model have been validated for a given gene, a first priority is given to gene-specific models with the highest AUROC. If there are no gene-specific models validated for a given gene, multi-gene models with the highest AUROC may be given next priority. Second, for major evidence categories (e.g., Protein Effect) that can be informed by multiple types of data and models, such as *in silico* and *experimental*, priority is given to the evidence criteria with the highest Sherloc point score. By combining both of these rules, we avoid double-counting similar or overlapping evidence and ensure that the weight of clinical and functional evidence are balanced appropriately within the classification framework.

### Concordance studies

#### Concordance between EMP models that use orthogonal data

To assess the reliability of the calibration steps for each model, we performed a concordance analysis between models that rely on orthogonal data for training. Models were categorized by the data used to train each model (column labeled Evidence Category in Table 1). Orthogonal models were defined as models that do not use similar or overlapping data for training and are therefore independent. For example, a model trained with population data (i.e., PFM) is considered orthogonal to models within the protein effect categories, which includes models trained with both experimental data and computationally derived data. Likewise, models within the experimental category are considered to be orthogonal to those in the computational category. A pairwise comparison was performed for each variant that had predictions derived from orthogonal models. We required that predictions for each model be within the high (≥ 95%) PPV or NPV bins. When two orthogonal models both predicted benign or both predicted pathogenic, they were considered concordant.

#### Concordance between EMP evidence criteria and internal variant classifications

To assess the reliability of EMP evidence criteria for variant classification, we identified the total set of variants at Invitae that were classified as pathogenic, likely pathogenic, likely benign or benign and had at least one modeling-related evidence criteria applied. Concordance was defined as true when the directionality of EMP evidence criteria (e.g., ≥ 1 pathogenic points) matched the variant classification (e.g., pathogenic or likely pathogenic), and false when the directionality (e.g., ≥ 1 benign points) did not match the variant classification (e.g., pathogenic or likely pathogenic). If a variant had both benign and pathogenic EMP evidence criteria placed, it was counted as both concordant and discordant. We discuss concordance results for models with at least 5 variants for a given bin, as a lower number can bias the comparison.

#### Prospective concordance between Invitae classifications and ClinVar submissions

To evaluate the reliability of variant classifications that were informed by EMP evidence, we compared *definitive* classifications (defined as P, LP, LB or B) made at Invitae with classifications that have been submitted to ClinVar by other clinical laboratories for the same variants. Because EMP models are often trained using ClinVar labels, our analysis design controls for circularity bias as follows (see also Figure S5). First, we limited the analysis to ClinVar submissions made after the latest iterative EMP model update described here (Q2 2022), which therefore could not have been used for EMP model training. These submissions were used to compose two groups of variants for concordance analyses: first, variants for which the first definitive classification (i.e., non-VUS) made to ClinVar was from an external lab and made after May 1, 2022 (stringent comparison group); for this group, only the first submission was used in analyses. Second, variants with ClinVar submissions made by external labs after May 1, 2022 irrespective of prior ClinVar records, but discarding any submissions made prior to this date (large comparison group). Note that the stringent group is contained within the large group. These two groups of ClinVar submissions were compared to Invitae definitive variant classifications made prior to May 1, 2022. We report both the concordance for all Invitae variant classifications meeting these criteria, as well as the subset of classifications informed by EMP evidence, resulting in four total concordance measurements. Concordance was defined as agreement between classifications. Pathogenic and Likely Pathogenic classifications were grouped as pathogenic while Benign and Likely Benign classifications were grouped as benign.

### Impact of EMP on variant classification rates for individuals undergoing genetic testing

To assess the impact of EMP evidence on the rates of classifications in a large-scale genetic testing laboratory, we first identified all variants with a definitive classification and at least one EMP evidence criteria applied. For these variants, we simulated the removal of all EMP evidence, and computed the new classification without EMP evidence. With this analysis, we were able to identify all variants in the Invitae database that would have been classified as VUS in the absence of EMP evidence; such variants were considered to have an *EMP-dependent classification*.

To measure the overall *clinical impact rate* of EMP evidence for individuals undergoing genetic testing, we counted the total number of individuals who received a genetic test at Invitae prior to October 1, 2023 and had at least one variant with an EMP-dependent classification. The clinical impact rate was computed across 12 main *clinical areas*: Hereditary Cancer, Preventive, Neurology, Pediatric Genetics, Cardiology, Metabolic Disorders, Immunology, Ophthalmology, Exome, Carrier, Hematology, Dermatology. Clinical areas were determined for each gene based on the commercial panel test(s) for which the gene is included (https://www.invitae.com/us/providers/test-catalog). As an example, *BRCA1* is associated with Hereditary Breast and Ovarian Cancer (HBOC) and included in multiple panel tests, all designed to provide information about a patient’s susceptibility to Hereditary Cancer. Therefore, a variant in *BRCA1* with an EMP-dependent classification was included in the Hereditary Cancer clinical impact rate. If a gene was included in panel tests from more than one clinical area, a single variant with an EMP-dependent classification was counted in each of the corresponding clinical areas. For example, *BRCA2* is included in both Hereditary Cancer and Pediatric Genetics, and the clinical impact from *BRCA2* is included in both of these clinical areas.

We also measured the clinical impact rate across the following REA groups: Ashkenazi Jewish, Asian, Black, Hispanic, Middle Eastern, Native American, Pacific Islander, Sephardic Jewish, White, Multiple/Mixed Ethnicity, Unknown/Other.

## Results

### The EMP enables robust increase in new evidence for clinical variant classification

With the Evidence Modeling Platform (EMP), nine types of evidence models were evaluated, calibrated and incorporated as evidence within our variant classification framework between August 1, 2019 and May 1, 2022 (Table 1). These models provide prediction scores for more than 10 million unique single nucleotide variants (SNVs) from more than 3,000 genes and encompass three major evidence categories for variant classification: population data, experimental, and computational. As of October 1, 2023, the end of the analysis period, 17 new evidence criteria (Table 2) have contributed to the classification of 864,364 unique variants observed in 1,052,843 patients.

### Prediction scores from orthogonal model types are highly concordant

Confidence in a clinical variant classification is greatly strengthened by agreement between orthogonal pieces of evidence. With the EMP, in this study, we developed predictive models that rely on (1) *population data*, (2) *computational data* (multiple sequence alignments, conservation scores, protein structural properties, physicochemical properties of amino acids, etc.) and (3) *experimental data*. Given the independence of the underlying data for these three categories of models, we performed a pairwise analysis between orthogonal models for each variant that had evidence placed from models in 2 or 3 of these categories. Results are shown in Figure 2A.

**Figure 2.**
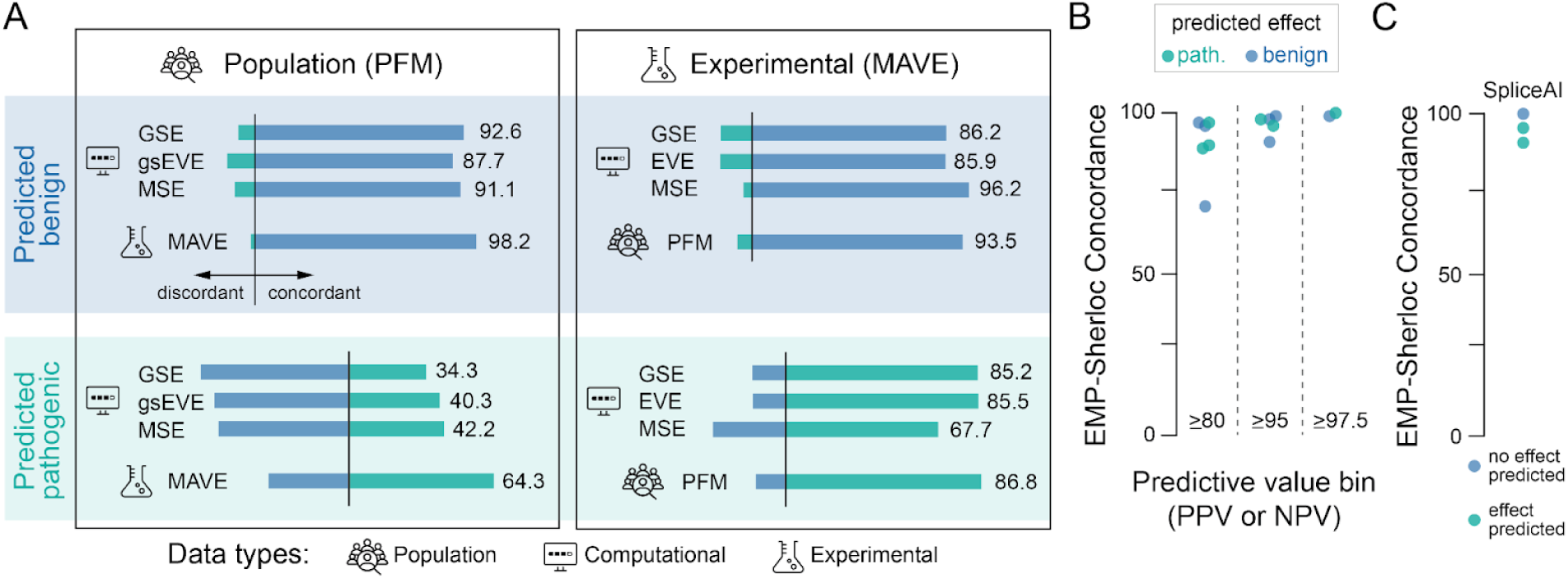
EMP Validation analysis. **A)** Concordance between orthogonal EMP evidence applied to the same variants. **B)** Concordance between EMP model evidence and final Invitae variant classifications as a function of calibrated predictive value bins. Each data point represents an EMP evidence criteria described in Table 2. One data point has been omitted due to small sample size (n=2). See Table 2 for the full dataset. **C)** Concordance between SpliceAI predictions and final Invitae variant classification.

High concordance (>85%) is observed between PFM benign evidence and all the other model types (for this analysis, DMA, DMX and DML models were combined into a single group referred to as MAVEs). In particular, PFM benign predictions are highly concordant with MAVE benign predictions. Much lower concordance (<65%) is observed on the pathogenic side between PFM and the other models. This makes sense considering a variant with relatively high frequency (> 1%) in the population is very likely benign, while the rare occurrence of a variant does not necessarily indicate that a variant is pathogenic. Due to this, PFM pathogenic scores were capped at 1 pathogenic point in Sherloc.

When we compare the two types of functional evidence, computational and experimental, predictions are also highly concordant (>85%) both on the pathogenic and benign sides with the exception of MAVE and MSE pathogenic predictions (67.7%). MSEs learn from structural information, such as FoldX and AlphaFold, while MAVEs learn from cellular effects. While MSEs evaluate the effect of mutations on structure, it does not capture all mechanisms whereby a missense mutation could be non-functional. For example, missense variants that impact protein-protein interactions would likely not score as pathogenic unless we have a structure of the protein complex.

### Calibrated EMP evidence criteria show high agreement with expert-reviewed classifications

To reach a definitive variant classification, multiple independent lines of evidence must agree;^10,30^ major lines of evidence include population data (e.g., low allele frequency in the general population), molecular effect (e.g., deleterious effect in a well-established functional study) and clinical observations of the variant (e.g., cosegregation with disease in multiple affected family members). Furthermore, each definitive classification made at Invitae is reviewed by a team of trained variant scientists and board-certified medical geneticists prior to being released to ordering providers and patients. Variants with conflicting lines of evidence, (i.e., some pathogenic and some benign) are either maintained as VUS or resolved by human experts using the available evidence. We therefore reasoned that concordance between the directionality of EMP evidence criteria (pathogenic or benign scores) and definitive classifications in our database represent an additional measure of accuracy for EMP evidence.

Overall, concordance was very high for benign (99.0%, N = 391,181 comparisons) and pathogenic (94.3%, N = 65,433 comparisons) EMP evidence criteria (see Table 2 for detailed results). Across all criteria, scores from the highest (≥97.5%) PV bins were the most concordant with Sherloc classifications (98.7 to 100% concordant), while those in the lowest (≥80%) PV bins had more variable concordance (71.0 to 97.4% concordant), with intermediate PV bins having high concordance (90.5 to 99.1%) (Figure 2B). SpliceAI predictions were highly concordant with Invitae classifications. While predicted splice gain or splice loss showed a 90.8% or 95.6% concordance with pathogenic predictions, the absence of splicing impact was 99.9% concordant with benign classifications (Figure 2C).

Because SpliceAI evidence represents a large proportion (79.7%) of the comparisons and could obscure results for other models, we also examined the overall concordance without SpliceAI observations, finding an overall concordance of 95.9% (N = 94,091 observations), with similar concordance for both benign (96.2%, N = 66, 451) and pathogenic (95.1%, N = 27,640) observations. Furthermore, we found that evidence criteria mapping to highly (≥95%) and very highly (≥97.5%) predictive bins had a concordance of 97.8% (N = 62,429) while the moderately predictive (≥80-95%) evidence criteria had a concordance of 92.1% (N = 31,662).

### Definitive classifications made with EMP evidence are highly concordant with prospective submissions to ClinVar

Utilizing the large repository of variant classification submissions within the ClinVar database, we compared definitive Invitae classifications made prior to May 1, 2022 to definitive classifications submitted by independent external labs to ClinVar after May 1, 2022 and before October 1, 2023. Using this start date allowed us to ensure the ClinVar variants used for evaluation were not also used for training EMP evidence models, avoiding the problem of circularity^42^ in this analysis.

We identified 652 variants in which the first definitive classification at Invitae was made prior to May 1, 2022 and the first definitive classification submitted by an external lab to ClinVar was after May 1, 2022. Importantly, Invitae’s submission to ClinVar for these variants was *not* the first submission so therefore did not influence the definitive classification from the other lab. With this set of variants, we found that 140 classifications at Invitae were informed by EMP evidence while 512 classifications were made without EMP evidence. The prospective concordance was 100% for variant classifications with EMP evidence and 99.2% for variant classifications without EMP evidence (Table S1). These results demonstrate a very high rate of independent, external corroboration for Invitae classifications. Significantly for this study, EMP modeling evidence did not degrade the quality of these classifications, further illustrating the quality of evidence derived from the EMP.

While this initial analysis provides the most stringent evaluation of prospective accuracy for Sherloc classifications with EMP evidence, eliminating the possibility that the EMP models were informed by the test set and that the test set was influenced by Invitae’s ClinVar submissions, the total number of comparisons (612) is quite small relative to the total number of variants receiving EMP evidence (864,364). In order to increase the number comparisons between ClinVar and Invitae’s classifications, we also identified a set of variants in which the first definitive classification at Invitae was made prior to May 1, 2022 and then compared these classifications to all new ClinVar submissions after May 1, 2022 (Group B, Supplemental Figure 5). This group may include multiple submissions from independent submitters for the same variant and did not require that the first ClinVar submission for a given variant occurred after May 1, 2022. This analysis allows the prospective evaluation of ClinVar submissions from multiple independent labs for the same variant. However, there are two important caveats: (1) it is possible that some of these variants may have been used for training EMP evidence if a definitive classification was submitted to ClinVar prior to May 1, 2022 and (2) it is possible that ClinVar submissions after May 1, 2022 may have been influenced by submissions prior to May 1, 2022. Nevertheless, this analysis yielded a substantial increase (40-fold) in the total number of comparisons (28,330). With this dataset, the concordance was 99.8% for variant classifications with EMP evidence and 99.7% for variant classifications without EMP evidence (Table S1).

### EMP evidence greatly increases the number of individuals with definitive classifications

In total, EMP evidence contributed to the classification of 864,364 unique variants, with 328,858 unique variants having a definitive classification that was dependent on the EMP evidence. As might be expected, the vast majority (>97%) of all EMP-dependent classifications were B/LB (319,226), while a much smaller fraction were P/LP (9,632). In terms of the impact of EMP-dependent classifications on patient results, we found that 30.6% (1,052,843) of all individuals undergoing genetic testing between September 1, 2013 and October 1, 2023 at Invitae received at least one variant classification supported by EMP evidence, while 15.9% of all individuals (545,805) would have received a VUS if the EMP evidence was not available for variant classification (95% (520,169) with one or more LB/B and 8% (41,876) with one or more LP/P classification).

The percentage of individuals who benefited from at least one EMP-dependent classification varied considerably by clinical area (Figure 3A). For example, at the high end, 48.3% (31,349/64,851) of patients tested for ophthalmology disease benefited from at least one EMP-dependent classification, while the fraction was much lower for Hematology at 5.6% (711/12,606) and Hereditary Cancer at 6.0% (112,433/1,856,961). Rare diseases within the pediatric genetics, metabolic and neurology clinical areas were in the middle with ∼20-30% of individuals benefiting from EMP-dependent classification.

**Figure 3:**
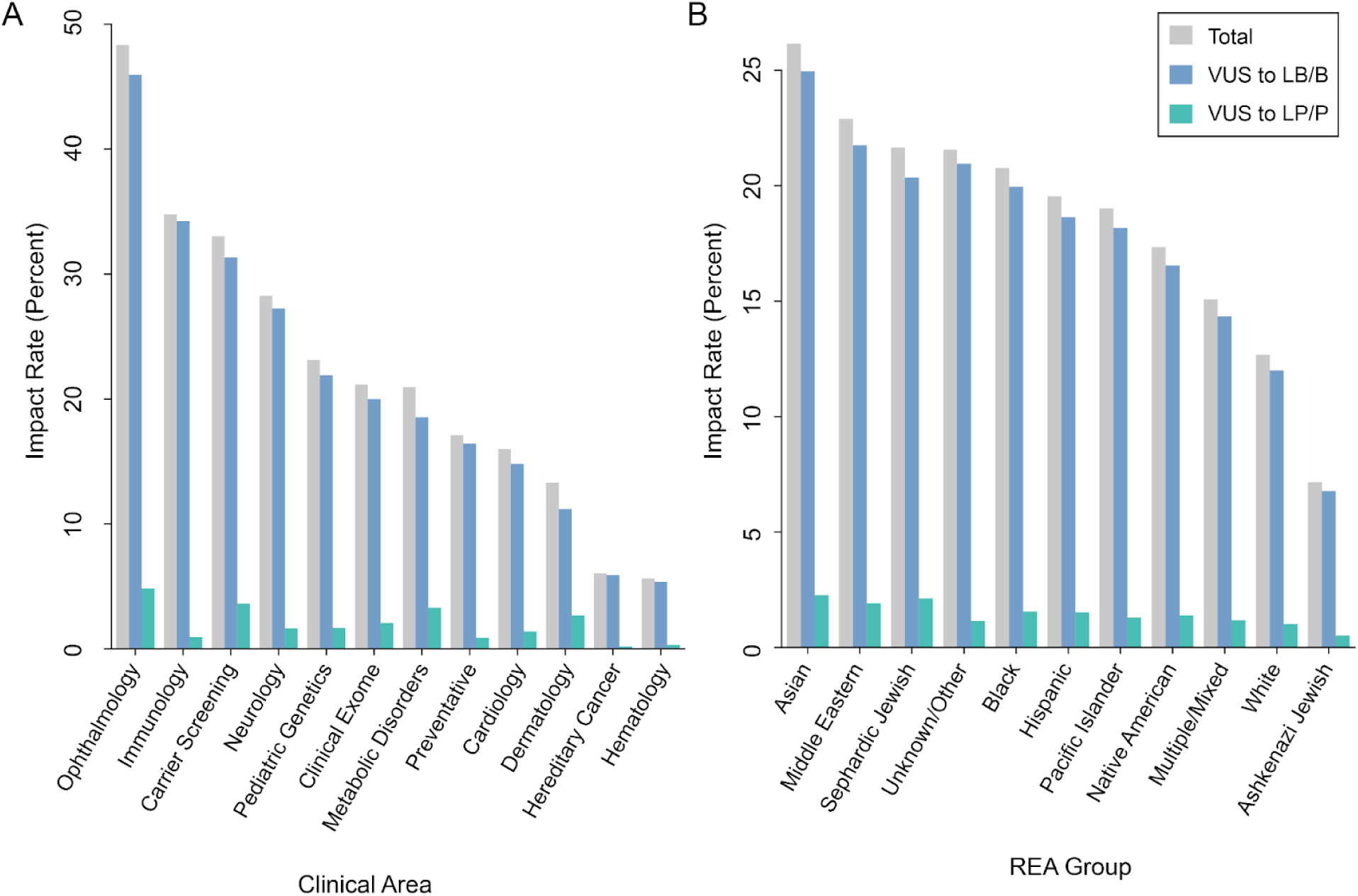
Impact of EMP evidence on variant classification. **A)** Patient level impact rate across clinical areas. **B)** Patient level impact rate across Race/Ethnicity/Ancestry (REA) groups.

When assessing the proportion of individuals who benefited from at least one *definitive EMP-dependent* classification across REA groups (Figure 3B), we found that significantly more individuals of Asian (26%), Middle Eastern (23%), Sephardic Jewish (22%), and Black (21.6%) backgrounds had at least one *definitive EMP-dependent* classification when compared to non-Hispanic white (12.7%) and Ashkenazi Jewish individuals (7.2%) (one-sided Fisher exact test, *p* < 10e^-20^).

## Discussion

The availability of data relevant to variant classification continues to grow at a rapid pace.^6,7,43–48^ While the expeditious increase of information characterizing a genetic variant offers promise, it is also more challenging to interpret due to its high volume and complexity. This underscores the need for an unbiased approach that can sift through the data at scale and provide evidence that can be used reliably in determining the pathogenicity of genetic variants. Here, through our Evidence Modeling Platform, we present a framework that optimizes the use of genetic and biological data for variant classification, leading to a reduction in VUS rates and increasing the utility of genetic testing.

Machine learning tools constitute powerful methods for identifying patterns and generating insights.^19–22,46,49,50^ While the integration of machine learning approaches is promising for variant classification at scale, it is essential that these approaches are robustly trained, evaluated and carefully chosen for the generation of accurate evidence. Some existing in silico predictors have been shown to be inaccurate,^51,52^ a problem that may stem from over-generalization of genome-wide model training. This approach inherently ignores the unique biological properties of individual genes. Additionally, meta-predictors combining multiple data types^53^ are vulnerable to circularity and can lead to overconfident classifications when combined with overlapping evidence in a variant classification framework.

Central to our Evidence Modeling Platform is the development of gene-specific, high-performing machine learning models using biologically relevant data. We use gene-specific validation tests with variants of known pathogenicity and ensure that these variants were not classified with data similar to the training data. This approach avoids circularity in the training process and over-inflated performance metrics. By training and evaluating variants in each gene separately, the EMP models are better able to learn specific biological and mechanistic features for each gene and provide more accurate predictions. Furthermore, we have implemented a selection framework where only the highest performing gene-specific models are considered for integration into our broader classification system and a calibration framework in which the weight of evidence being given for each variant is calibrated based on the accuracy metric of the gene-specific validation. The platform’s systematic and stringent design ensures reliability, ease of use, and iterative improvement.

Our findings demonstrate the quality and high performance of the EMP variant predictions when used as evidence for variant classification. First, variant pathogenicity predictions from models trained on orthogonal data show good concordance overall, suggesting a high degree of confidence in the predictions from high-performing models within EMP. Secondly, the direction of EMP evidence criteria and our expert-curated definitive variant classifications also showed high concordance for both benign (%) and pathogenic (%) variants, adding further confidence in the EMP-generated evidence. Lastly, a prospective concordance analysis between EMP-informed variant classifications and submissions to ClinVar by external labs evaluating the same variant showed perfect concordance (100%). By comparison, a 2018 analysis of inter-lab discordances across all of Clinvar observed a concordance of 77.8% when two or more labs submitted classifications for the same variant.^54^ Altogether, these validation studies show that EMP can generate highly accurate and consistent evidence for variant classification.

To illustrate the impact of EMP in resolving VUS, we analyzed data from over 3 million individuals tested in our lab over a ten-year period. Over 16% (545,805) of these individuals received a definitive variant classification (P, LP, LB, B) that would have remained a VUS without EMP evidence. The vast majority of these individuals (520,169; 95.3% of the total) received one or more LB/B classifications, while a much smaller proportion (41,876; 7.7% of the total) received at least one LP/P classification. Of note, some individuals received both LB/B and P/LP variants which is why the percentages don’t add up to 100%. These results align with the expectations that most rare variants are not disease-causing^31,55,56^ and ∼7-20% of individuals undergoing clinical genetic testing have one or more pathogenic variants.^14,57,58^ The benefit of EMP-dependent variant classification varied across clinical areas, with some, like ophthalmology, seeing greater benefit compared to others, such as hematology and hereditary cancer. This can largely be explained by the different proportion of VUS across clinical areas.^14^ Overall, 5-50% of individuals in each clinical area benefited from an EMP-dependent classification (Figure 3A).

Historically, non-white populations have had higher VUS rates, primarily due to their underrepresentation in genetic databases (e.g. gnomAD, UK Biobank), genetic testing cohorts^59,60^ and clinical trials.^61,62^ While some types of EMP evidence, like PFM, are potentially limited by underrepresentation of certain REA groups, other evidence types leverage data based on core principles of protein biology—such as structure, impact on cellular phenotype, and sequence conservation—that offer biological and functional evidence that are less prone to representation biases. With the EMP and across all model types, our results demonstrate a much greater reduction in the number of VUS for historically underrepresented populations, specifically those of reported Asian, Middle Eastern, Sephardic Jewish, and Black descent.

These findings highlight the potential that thoughtfully trained ML models can help close the gap in definitive results across these groups, contributing to a more equitable genetic testing landscape.

To ensure high quality evidence, the EMP focuses on gene specific models whenever possible, a strategy that can be limited by the number of known pathogenic and benign variants that can be used to train models for less well-understood genes. Multi-gene models offer the opportunity to combine labels across multiple genes but it is sometimes at the detriment of the quality of the predictions. In such scenarios, disease-specific approaches could increase the number of labels for training while mitigating quality concerns.^63^ Unsupervised learning methods could also be key in finding patterns associated with disease risk.^64,65^ Additionally, initial EMP models have focused primarily on single nucleotide variants (i.e., missense, nonsense, and synonymous).

While this class of variants constitutes the largest proportion of VUS, other variant types, such as copy number variations^66^ or tandem repeats^67^ among others are disease causing as well. Modeling additional variant types, including those residing in non-coding regions, is possible and represents an important next step for EMP development.

In conclusion, the rapid expansion of data relevant to variant classification presents both opportunities and challenges. The development of the Evidence Modeling Platform (EMP) represents a significant advancement in our ability to harness these new data in a clinically responsible way. By standardizing the generation and evaluation of gene-specific, expert-informed machine learning models, the EMP substantially increases rates of definitive classifications while maintaining high quality. The platform’s design also facilitates the retraining of existing models, development of new models and serving of updated evidence with minimum overhead. Because the EMP model outputs are rooted in key evidence categories and explainable, they can serve as inputs into other classification systems, in particular as the field moves towards a more quantitative assessment of variant pathogenicity.^13,68–70^ Lastly, while not obviating the need for increased representation in genetic databases, we demonstrate that carefully designed modeling approaches, like the EMP, have the potential to increase equity and narrow the gap in generating definitive genetic results for individuals from diverse REA groups.

## Data and code availability

Variant classifications are submitted to Clinvar by Labcorp (formerly Invitae). Code for SpliceAI improvements and modifications are available https://github.com/invitae/SpliceAI. The remaining datasets and code supporting the current study have not been deposited in a public repository because they are proprietary.

