## Supplementary Material for "The impact of systematized generation, evaluation, and incorporation of machine learning algorithms for clinical variant classification"

### Supplemental Materials and Methods

Details given in the following sections correspond to specific versions of the models used within the time period of the described study (between September 1, 2013 and October 1, 2023). Within the EMP framework, models are updated and while the feature categories will remain, methods might change. For example, more recent data is made available and will impact the model version specific description.

#### Population Frequency Modeling (PFM)

*Features.* Data from gnomAD (v2.1.1)[1] including ancestry specific allele counts, allele numbers, and gene-level constraint estimates (e.g., LOEUF) were collected for single nucleotide variants in genes with at least one strong disease association according to our internal genetics knowledge base.

*Model Training.* We identified a total of 33,382 labels (pathogenic = 15,234; benign = 18,148) available across 823 genes. The overall AUROC for this model was 0.92, with 232 genes flagged for poor calibration (Brier score > 0.15) or poor discrimination (AUROC <0.8). In total, PFM v1 output scores from 591 genes with AUROC $\geq$ 0.8 were standardized and incorporated into the population data category of Sherloc.

*Calibration.* For the point score bins, we assigned 5B (NPV>99%), 3B (NPV>95-99%), 1B (NPV>80-95%) and 1P (PPV>95%) to a new class of evidence criteria (Table 2). By incorporating the PFM v1 into our clinical variant classification workflow, we have applied population evidence to 4x more variants than other methods developed to establish population frequency thresholds,[2] leading to more than 15,000 variants reclassified and impacting more than 50,000 patients at the time of implementation and estimated to provide ongoing definitive classification rate of 2.5% for variants new to Invitae.

#### SpliceAI

*Features.* The SpliceAI model was previously developed by[3] and uses transcript sequences to predict the probability of a splice change given a variant.

*Validation.* The sensitivity of SpliceAI was assessed at Invitae using a set of 1,236 true positive variants with observed splicing shown through *in vitro* or *in vivo* RNA analysis or *in vitro* splicing assays. SpliceAI scores above or equal to 0.2 were considered to be positive predictions by SpliceAI. Overall, spliceAI yielded 91.34% sensitivity (1129/1236) to detect true splicing events. The specificity of SpliceAI was assessed using a set of 81 true negative variants that were not observed to impact splicing by RNA analysis or *in vitro* studies. SpliceAI scores less than 0.2 were considered to have a negative splicing prediction. The specificity of SpliceAI was 79% (64/81).

*Calibration.* We used a modified version of SpliceAI allowing batch processing of variants in order to generate predictions (https://github.com/invitae/SpliceAI).
Variants with SpliceAI predictions ≥0.2 were given 1 pathogenic point. Variants for which predictions were $<$ 0.2 threshold were given 1 benign point.

**Gene specific engine (GSE)**
*Features.* GSEs leverage patterns in sequence coevolution (developed by Invitae), physico-chemical properties, mobility, protein conservation metrics, context of molecular variants within genes.

*Training.* GSEs were trained on 957 genes with a minimum of 8 pathogenic and 8 benign variants. In total, 664 genes were performant enough (AUROC>=0.8) to be considered for integration.

*Calibration.* We assign evidence to the variants using PPV/NPV thresholds. 95% PPV/NPV results in 2 PP or BP, respectively and 80% PPV/NPV results in 1 PP or BP, respectively.

**Molecular stability engine (MSE)**
*Features.* Molecular Stability Engines (MSEs) are an evidence model that predicts the effect of missense mutations on protein stability. To do this, MSEs use protein structure data, introduce variants into these structures and calculate predictions from the changes in protein stability from the variant structures. In addition to this change in stability prediction (ΔΔG) or the variant in question, the model includes a few other engineered features to improve the quality of the model.

- ΔΔG value = predicted change in stability from FoldX calculations
- ΔΔG position metric = median ΔΔG value of all the mutations at a particular protein position
- FoldX median substitution metric = similar to a BLOSUM score, this feature is a mutation-specific (i.e. there is a value for each kind of missense mutation, such as A->G, W-L, etc.) median ΔΔG value over all observations of this mutation in the FoldX calculation database (over 2M mutations)
- ΔΔG window metric = moving window median ΔΔG position metric value using a Gaussian function and a window of 5

*Training.* MSEs are trained on individual transcripts. Only transcripts containing at least 5 pathogenic and 5 benign labels in positions with ΔΔG data (which is a subset of transcript positions as not all positions have structures). Labels are determined by the EMP labeling pipeline, which uses a list of approved submitters to consolidate interpretations from ClinVar, and also includes population information and other data. The labels for training were generated using ClinVar data from prior to March 2, 2021. From the labels, 20% of labels are withheld from training and used for test set evaluation to check model generalizability.

*Calibration.* We assign evidence to the variants using PPV/NPV thresholds. 95% PPV/NPV results in 2 PP or BP, respectively and 80% PPV/NPV results in 1 PP or BP, respectively.

**Deep mutational assays (DMA) and Combined deep mutational assays (DMX)**
*Features.* DMA models were trained with multiplexed assays of variant effects (MAVE) data for 49 genes curated from 22 publications[4–26]. DMX was trained with functional assay data from three separate publications focused on the *TP53* gene[5,6,17] that were combined into one model.

*Training.* DMAs and DMX were trained at the transcript level. Labels are determined by the EMP labeling pipeline, which uses a list of approved submitters to consolidate interpretations from ClinVar, and also includes population information and other data. The labels for training were generated using ClinVar data from prior to March 2, 2021. From the labels, 20% of labels are withheld from training and used for test set evaluation to check model generalizability.

*Calibration.* We assign evidence to the variants using PPV/NPV thresholds. For DMA models, 95% PPV/NPV results in 2 PP or BP, respectively and 80% PPV/NPV results in 1 PP or BP, respectively. For the DMX model, 97.5% PPV/NPV results in 2.5 PP or BP, respectively and 80% PPV/NPV results in 1 PP or BP, respectively.

**Deep mutational learning (DML)**

*Features.* DML models were trained with MAVE data generated in our laboratory for 44 genes[26]. The input to DML models is single cell RNA sequencing (scRNA-seq) data generated by Invitae's cellular evidence platform lab. To assess the effect of variants, we measure the gene expression of single cells with a variant of interest in a target gene.

*Training.* The data undergoes standard scRNA-seq preprocessing methods and dimensionality reduction. The resulting latent space matrix is fed into a classifier that trains the data on ClinVar labels and predicts the probability of pathogenicity of the variants.

*Calibration.* We assign evidence to the variants using PPV/NPV thresholds. 95% PPV/NPV results in 2 PP or BP, respectively and 80% PPV/NPV results in 1 PP or BP, respectively.

**Evolutionary model of variant effects (gsEVE and mgEVE)**

Evolutionary model of variant effect (EVE) is a published unsupervised deep generative learning model that predicts the pathogenicity of missense variants based on multiple sequence alignments.[27] EVE models use a deep learning strategy called Variational Autoencoders (VAEs) to learn about the underlying latent representation of sequence space.

*Features.* The input for this published algorithm is a collection of protein sequences from a variety of organisms - called a multiple sequence alignment (MSA) - that represents the evolutionary history of the protein and provides information on the sequence constraints on an amino acid in a protein sequence.[27]

*Training.* In order to incorporate the published EVE model into the Sherloc evidence framework, we trained a gene-specific model (gsEVE) and a multi-gene model (mgEVE) with the EVE prediction scores as features. For genes with $\geq$ 8 pathogenic and $\geq$ 8 benign labels, we utilized the gene-specific training and validation approach described in the primary methods. To expand the scope of the EVE model for genes with $\leq$ 8 pathogenic or $\leq$ 8 benign labels, we aggregated 64,304 (pathogenic = 25,855; benign = 38,449) labels across 3333 genes and trained a single model. The overall AUROC for this multi-gene model was 0.87.

*Calibration.* For gsEVE, we then assigned evidence according to the following thresholds: 95% PPV/NPV results in 2 PP or BP, respectively, 80% PPV/NPV results in 1 PP or BP, respectively. We quality control transcripts such that the performance overall reaches an AUROC of 0.8, resulting in transcript-specific EVE models for over 700 transcripts. These transcript-specific models are able to assign evidence for more than 1 million variants. For mgEVE, we defined a single threshold for 80% PPV/NPV since we were unable to calibrate PPV/NPV at the gene-level.

**The rationale for gene specific validation**

The overall performance of a given model differs from gene to gene (Supplemental Figure 4). While a model type (see Table 1) may perform well for a specific gene or group of genes based on the features of the gene(s), it may not perform well for others.

A model that is able to discriminate well between pathogenic and benign variants will receive higher weights for its predictions in the pipeline for variant classification. In contrast, a model that shows poor discrimination between pathogenic and benign variants in a specific gene, will not be used in the variant classification pipeline.

This gene specific validation of each model is thus critical when considering the integration of prediction from a model into our variant classification system.

### Supplemental figures and tables

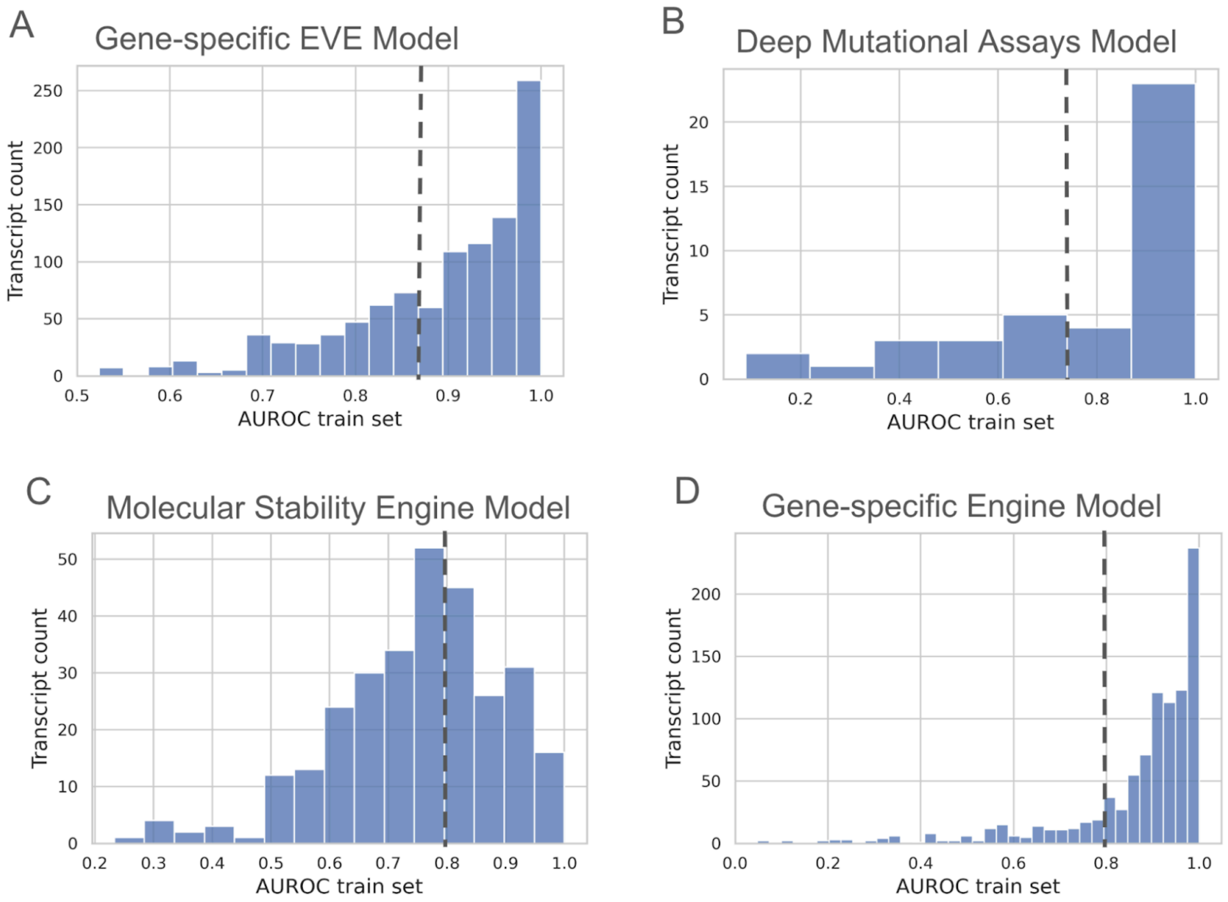


***Figure S1. Transcript performance distribution for each model type.*** *The area under the receiver operator curve (AUROC) performance metric is calculated for each transcript and is shown on the X axis from 0.0 to 1.0. The number of transcripts with a given AUROC is shown on the Y axis. The dotted lines indicate the threshold (0.8) at which a model is excluded from the remaining steps in the EMP pipeline. Model types shown include (A) Gene-specific Evolutionary Variant Effect (EVE) model, (B) Deep Mutational Assays (DMA) model, (C) Molecular Stability Engine (MSE) model, and (D) Gene-specific Engine (GSE) model.*

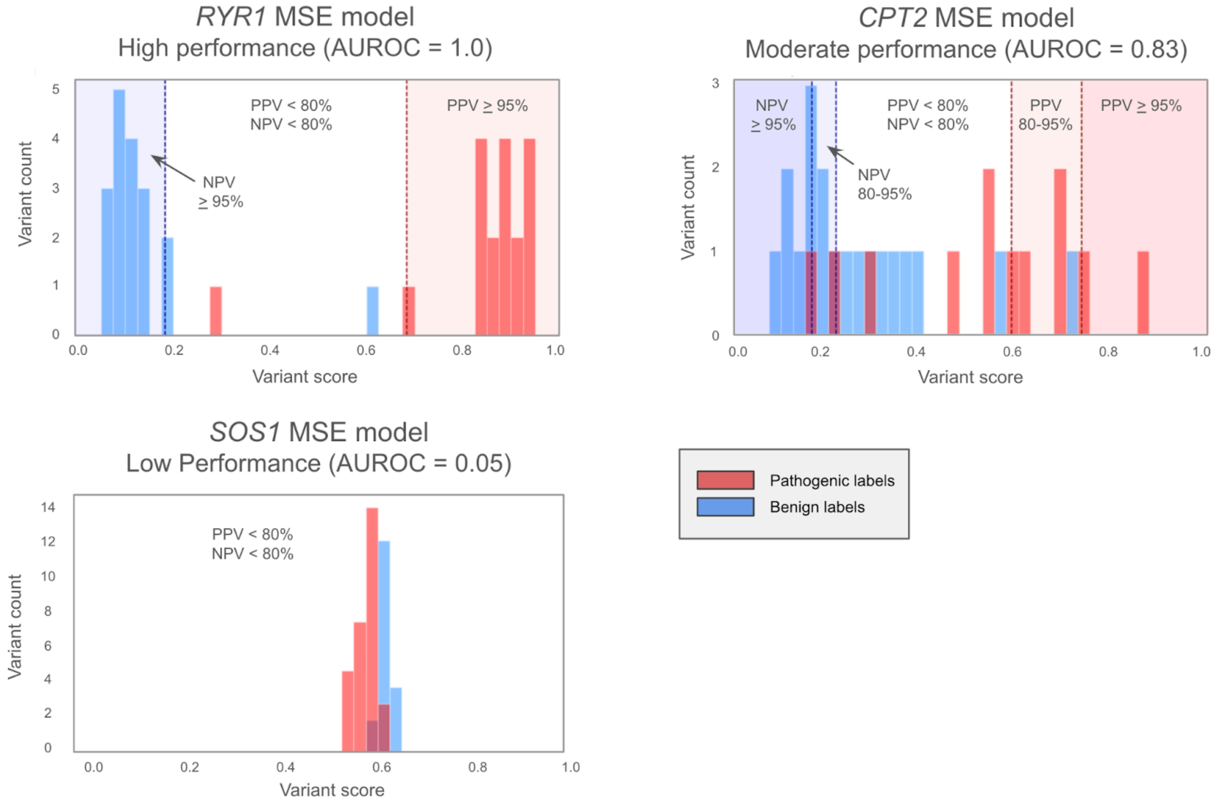


***Figure S2. Calibration examples of gene-specific models with high, moderate and low performance metrics.*** *These plots show the variant score distribution of known pathogenic and benign variants (labels) for three gene-specific (RYR1, CPT2, SOS1) molecular stability engine (MSE) models with high (AUROC = 1.0), moderate (AUROC = 0.83) and low (AUROC = 0.05) performance metrics. During the calibration step described in the methods, positive predictive value (PPV) and negative predictive predictive value (NPV) bins were determined and shown in red or blue, respectively. Variant score is shown on the X axis and variant count is shown on Y axis. Pathogenic labels are shown in red and benign labels are shown in Blue.*

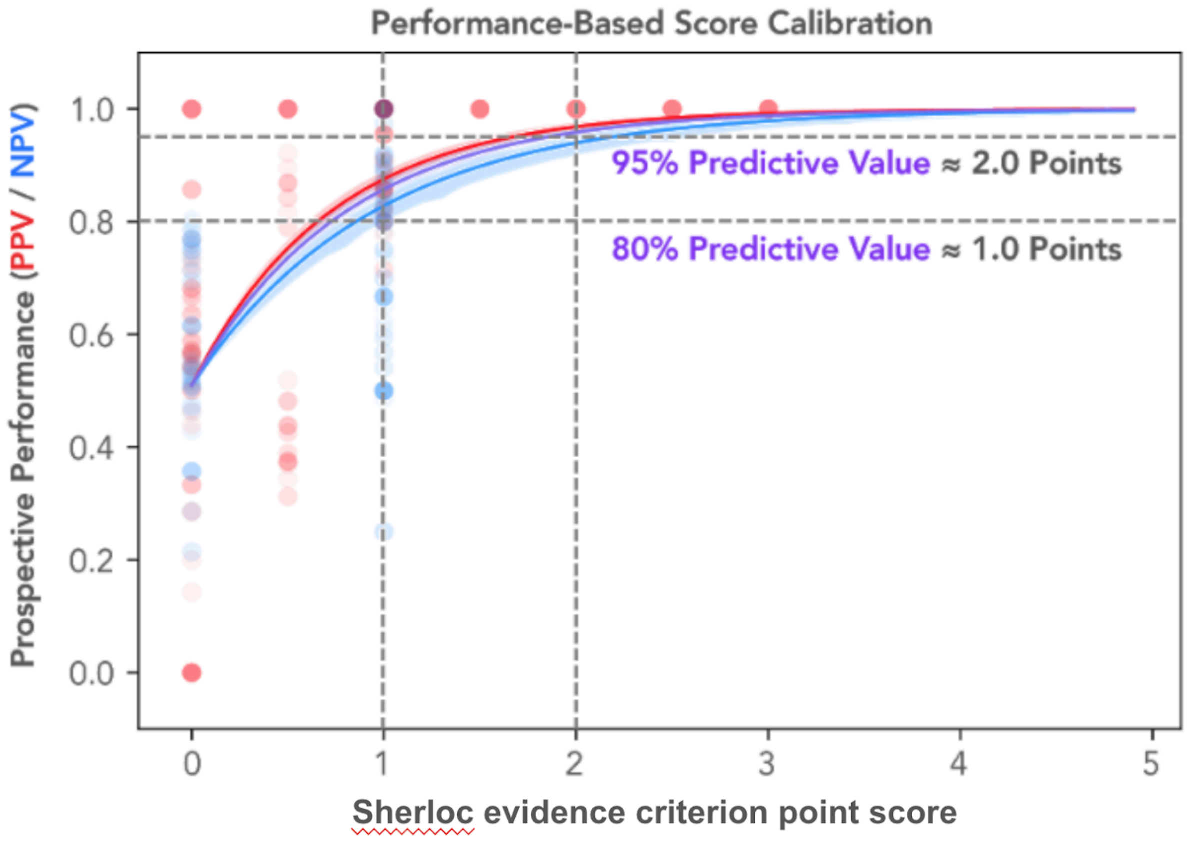


***Figure S3. Calibration of Sherloc evidence point scores.*** *Positive predictive values (PPV), for pathogenic criteria, and negative predictive values (NPV), for benign criteria were calculated for all available Sherloc evidence criteria. Each dot is a single evidence criteria; red dots denote a Pathogenic criteria and blue dots denote a benign criteria. The evidence criterion point score (0 - 5) is provided on the X axis and the calculated PPV (red) or NPV (blue) is plotted on the Y axis. Based on the regression curves (PPV: red, NPV: blue, combined: purple), we estimate that 2 pathogenic or 2 benign points meet a 95% predictive value threshold, while 1 pathogenic or 1 benign point meets a 80% predictive value threshold.*

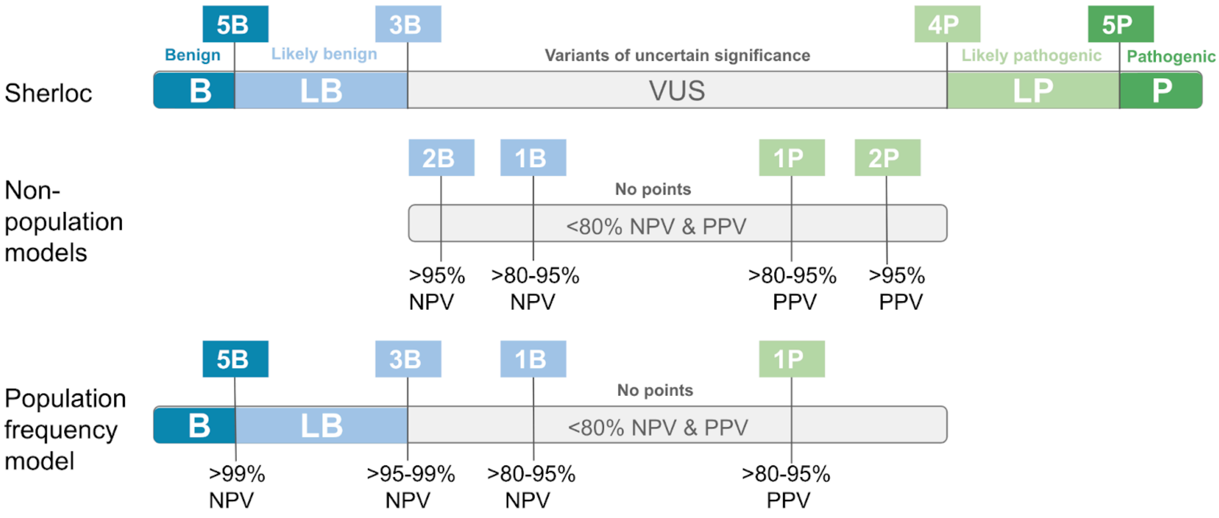

***Figure S4. An illustration for how predictive value (PV) bins are assigned points scores in the Sherloc evidence framework.*** *Top diagram, the point score thresholds for Benign (5B, benign points), Likely benign (3B, benign points), Likely pathogenic (4P, pathogenic points) and Pathogenic (5P, pathogenic points). Variants of uncertain significance (VUS) are variants that do not reach one of the more definitive thresholds or reach both pathogenic and benign thresholds. Middle diagram, the point scores assigned for NPV/PPV bins for non-population frequency models (e.g. EVE, MSE, GSE, DMA, DML). Lower diagram, the point scores assigned for NPV/PPV bins for the population frequency model (PFM).*

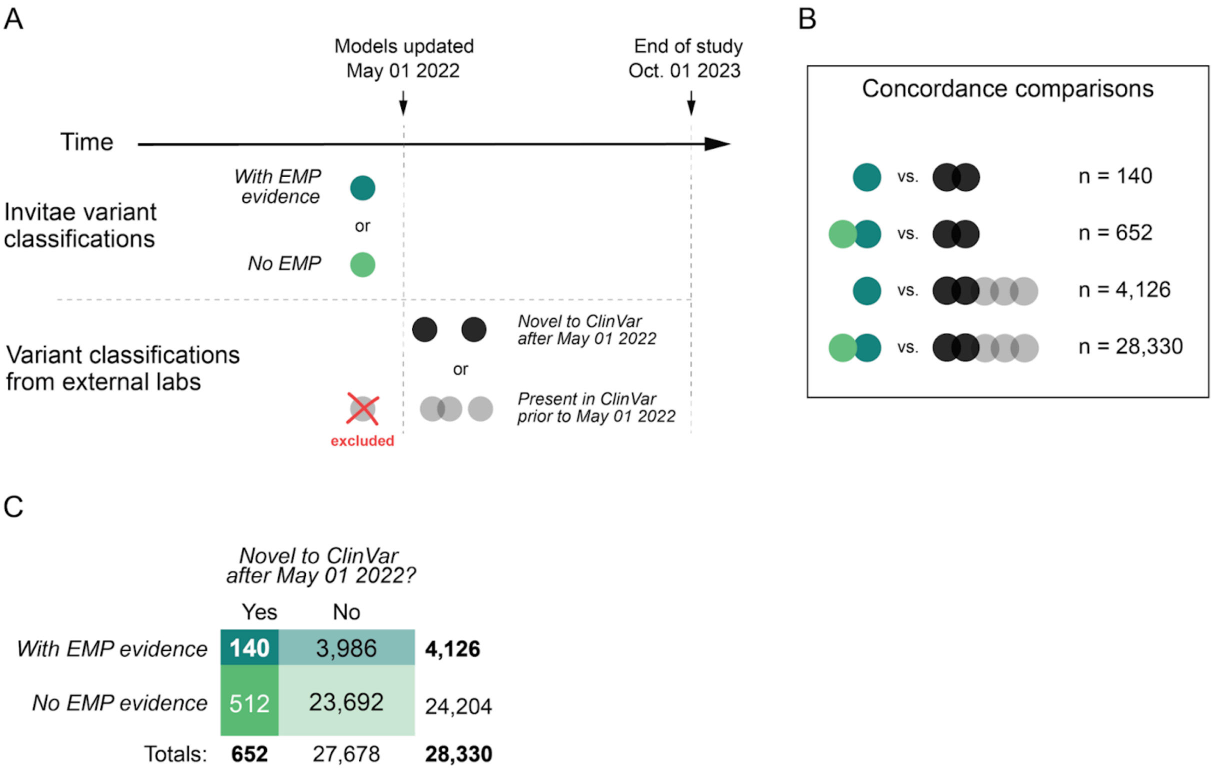


***Figure S5. An illustration of the Clinvar prospective concordance analysis.*** *A) All EMP models described in this study were trained, validated and integrated into Sherloc variant classification framework prior to Q2, 2022. All Invitae variant classifications made prior to this date were identified and grouped into two categories: with EMP evidence (dark green dot) or without EMP evidence (light green dot). Comparisons were made between the Invitae classifications and Clinvar submissions made after May 01, 2022 and before Oct 01, 2023. We identified two groups of variants with Clinvar submissions: variants in which all Clinvar submissions were made after May 01, 2022 are shown in black, variants in which at least one submission was made prior to May 01, 2022 are shown in grey. Notably, we excluded the submission prior to May 01, 2022 from the concordance analysis. Although, it is possible that a variant with a submission prior to May 01, 2022 was used as a label for training some or all of the EMP models. B) We performed four concordance comparisons with an increasing number of variant submissions. C) Rates of concordance for each of four comparisons.*

| | First submission to ClinVar (stringent) | All new laboratory submissions (inclusive) |
|---|---|---|
| Variants classified by Invitae without EMP evidence | 99.2% (n=512) | 99.7% (n=24,204) |
| Variants classified by Invitae with EMP evidence | 100% (n=140) | 99.8% (n=4,126) |

***Table S1. Concordance between variants classified at Invitae and Clinvar submissions.***

http://dx.doi.org/10.1016/j.ajhg.2020.05.015
